# Clinical features of imported cases of coronavirus disease 2019 in Tibetan patients in the Plateau area

**DOI:** 10.1101/2020.03.09.20033126

**Authors:** Yu Lei, Xiaobo Huang, Bamu SiLang, YunPing Lan, Jianli Lu, Fan Zeng

## Abstract

Coronavirus disease 2019 (COVID-19), caused by SARS-CoV-2, has rapidly spread throughout China, but the clinical characteristics of Tibetan patients living in the Qinghai-Tibetan plateau are unknown. We aimed to investigate the epidemiological, clinical, laboratory and radiological characteristics of these patients. We included 67 Tibetan patients with confirmed SARS-CoV-2 infection. The patients were divided into two groups based on the presence of clinical symptoms at admission, with 31 and 36 patients in the symptomatic and asymptomatic groups, respectively. The epidemiological, clinical, laboratory and radiological characteristics were extracted and analysed. No patient had a history of exposure to COVID-19 patients from Wuhan or had travelled to Wuhan. The mean age of Tibetan patients was 39.3 years and 59% of the patients were male. Seven patients presented with fever on admission and lymphocytopenia was present in 20 patients. 47 patients had abnormal chest CTs at admission instead of stating that 20 were unchanged. Lactate dehydrogenase levels were increased in 31 patients. Seven patients progressed to severe COVID-19; however, after treatment, their condition was stable. No patients died. Of the 36 asymptomatic patients, the mean age was younger than the symptomatic group (34.4±17.3vs 44.9±18.1 years, P=0.02). Lymphocyte count and prealbumin levels were higher in the asymptomatic group than the group with clinical symptoms (1.6±0.5 vs 1.3±0.6 and 241.8±68.2 vs 191.9±60.3, respectively; P<0.05). Imported cases of COVID-19 in Tibetan patients were generally mild in this high-altitude area. Absence of fever or radiologic abnormalities on initial presentation were common.

## Introduction

Corona virus disease 2019 (COVID-19) has rapidly spread from Wuhan to other areas of China and has now become a global threat. At the time of writing on March 2^nd^ 2020, COVID-19 cases have been confirmed in 92 countries, with more than 100,000 cases globally. Wuhan is thought to be the site of earliest COVID-19 occurrence, and cases further afield were infected by SARS-CoV-2 carriers from Wuhan. In particular, the mortality of patients in Wuhan was higher than in any other city in China, at 4.3% compared with 0.8% in the rest of mainland China(updated data available at https://wp.m.163.com/163/page/news/virus_report/index.html?spss=feed&&spssid=a64bed4d89174914f9792895db5b15e8&spsw=1). The clinical characteristics and outcome of patients seem to be different between Wuhan and other areas. Despite the publication of many articles regarding the clinical features of COVID-19 patients, most of these patients were considered in the context of Wuhan. Daofu, located within the Qinghai-Tibetan Plateau at an altitude of more than 3000 m, is a low-income county in Sichuan Province, China. There are more than 3000 Tibetans living here. Of those becoming infected, none of them had travelled to Wuhan or had a history of exposure to COVID-19 cases from Wuhan. The clinical characteristics in Tibetans living in these highlands were unclear. Thus, herein we provide an analysis of data from these patients describing epidemiological, clinical laboratory and radiological characteristics, treatment and outcome.

## Methods

### Patient enrollment and data collection

Daofu People’s Hospital was the only hospital that could accept COVID-19 patients. Since the first patient with confirmed COVID-19 on Feb 4^th^, the local Centre for Disease Control and Prevention (CDC) collected nasal and pharyngeal swabs or blood to detect viral nucleic acids by real-time PCR from more than 8000 people living in Daofu, with and without clinical symptoms. Those with positive samples were taken to Daofu People’s Hospital for further tests. We enrolled all patients who had been confirmed as SARS-CoV-2 carriers from Jan4^th^–Feb 28^th^, according to a rapid advice guideline by the Wuhan university Novel Coronavirus Management and Research Team ^1^ The clinical outcomes were monitored until March 5^th^. This study was approved by the institutional ethics board of Sichuan Hospital.

Epidemiological, clinical laboratory and radiological characteristics, chronic medical histories, clinical symptoms, treatment and outcome data were obtained from electronic medical records and analysed by two independent researchers. When missing or uncertain records were encountered, the researchers communicated directly with patients or their families to collect and clarify the relevant data. The date of disease onset was defined as the day when symptoms were first noticed or the day when the real-time PCR test for nucleic acid in respiratory or blood samples from asymptomatic patients was positive. The patients were then divided into two groups based on the presence of clinical symptoms at time of admission. The symptomtic group was defined as those patients with any clinical symptoms such as fever, cough and headache. According to the diagnostic and treatment guidelines for COVID-19 issued by the Chinese National Health Committee, severe COVID-19 was defined as the occurrence of either one of the following criteria: respiratory distress with respiratory frequency ⩾30/min; oxygenation index (artery partial pressure of oxygen/inspired oxygen fraction, PaO_2_/FiO_2_) ⩽300mmHg. Importantly, oxygenation index should be corrected if the local altitude is higher than 1000 m using the corrector formula: PaO_2_/FiO_2_*atmospheric pressure/760.

### Laboratory testing

Patient nasal and pharyngeal swabs or blood samples were collected for detection of SARS-CoV-2 viral nucleic acid using real-time PCR assay. Laboratory confirmation of SARS-CoV-2 was performed by the local CDC as previously described ^1^.

### Statistical analysis

Categorical variables were summarized as frequencies and percentages. Continuous variables were expressed as median + standard deviation (SD) or inter-quartile range (IQR). Continuous variables were compared using Student’s *t*-test and the Mann-Whitney test. The chi-squared and Fisher’s tests were used for the frequencies of categorical variables. All statistical analyses were performed using SPSS software (version 24, IBM, Armonk, NY). *P*-values less than 0.05 were considered to be statistically significant.

## Results

### Clinical findings

A total of 67 patients diagnosed with COVID-19 were included in this study. All of them were Tibetans living in Qinghai-Tibetan plateau Twenty-six were members of one family, 34 had a history of attending a funeral or temple; however, none had visited Wuhan or had contact with Wuhan residents. The first patient to be diagnosed with SARS-CoV-2 infection had travelled to Chengdu,the city of westchina., 10 days before onset of symptoms but denied any contact with COVID-19 patients. The clinical characteristics of the patients are shown in Table 1.2. Thirty-nine of 67 patients (58.2%)were male, with a median age of 39.3 years. The youngest patient was 3 years old, with his family all confirmed as SARS-CoV-2-positive. Twenty (29.9%)of the total cohort had chronic diseases. Fever was present in only seven patients (10%) on admission and developed in a further nine (24%) during hospitalization. Thirty-six patients (54%) showed no clinical symptoms when they were admitted to hospital. The remaining 31 patients (46%) presented with clinical symptoms, of which cough was the most common (13/67, 19%). Other symptoms included fatigue, headache, muscle ache and dizziness. Of note, the asymptomatic group was significantly younger than the symptomatic group, with median ages of 34 and 44 years (P=0.02), respectively.

**Table 1.**
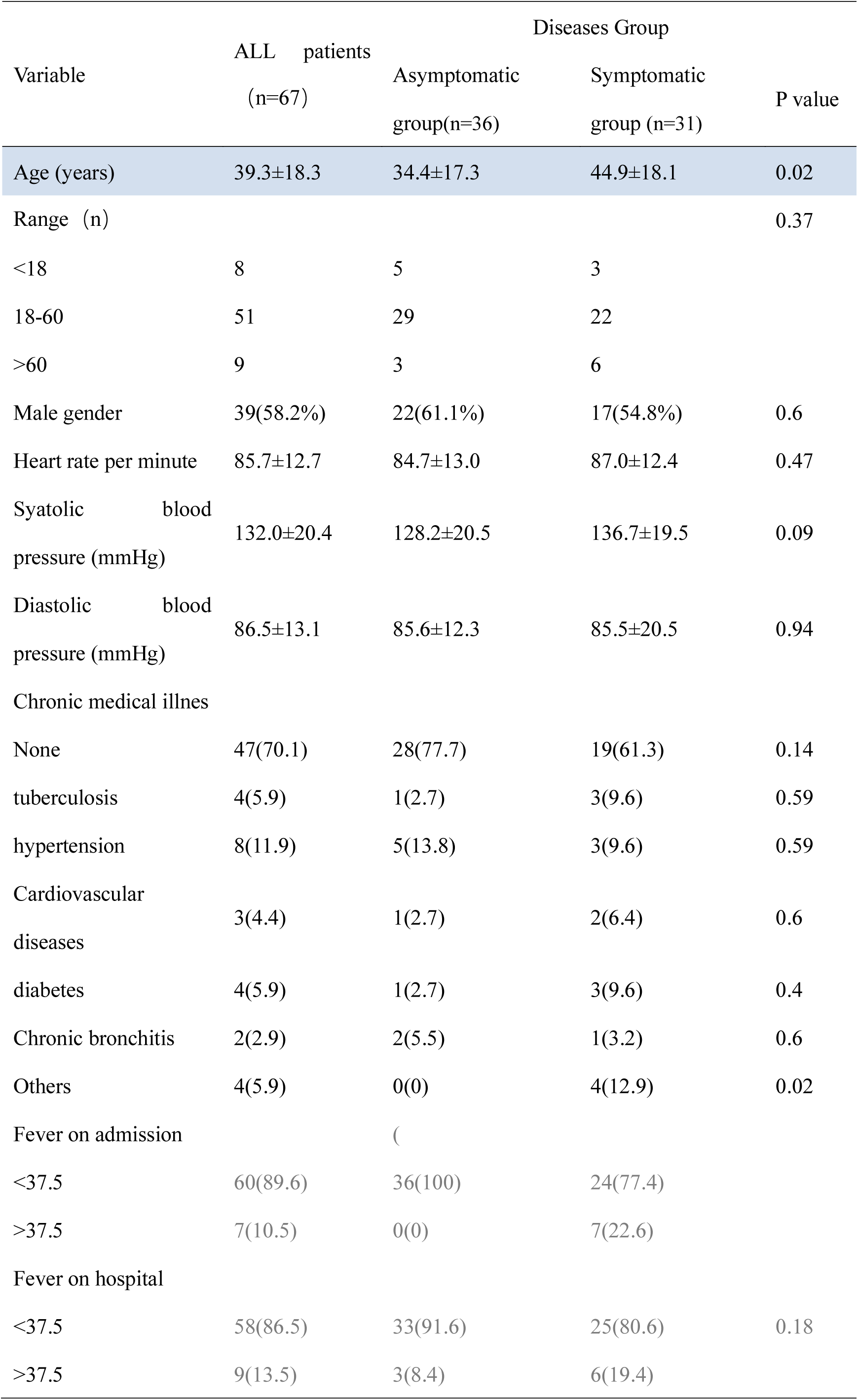
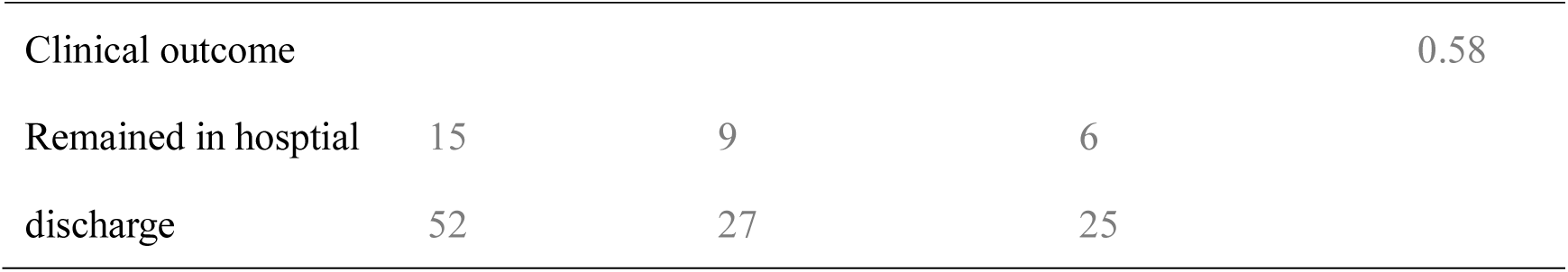
Baseline characteristics on admission and clinical outcomes of patients with COVID-19

**Tbale 2.** Clinical Symptoms of patients with COVID-19 at admission

| Symptoms at admission | Number of patients |
| --- | --- |
| None | 36(53.7%) |
| Fever | 7(10.4%) |
| Fatigue | 5(7.4%) |
| Palpitationri | 1(1.5%) |
| Headache or dizzy | 9(13.4%) |
| dyspnoea | 2(2.9%) |
| Sore throat | 1(1.5%) |
| Muscle ache | 3(4.4%) |
| Cough | 13(19.4%) |
| Diarrhoea | 1(1.5%) |

### Radiologic and laboratory finds

All patients consented to CT scans on admission. Forty-seven patients (70%) had abnormal results, with 21 and 26 patients in the symptomatic and asymptomatic groups, respectively, of which ground glass opacity was the most common pattern and bilateral onset was the main manifestation. Chest CT imaging changed in 21 of the patients without clinical symptoms. In five patients with clinical symptoms, chest CT imaging showed no change. Liver injury was found in 24 of 67 patients (36%), but the injury was very light. Increases in alanine aminotransferase or aspartate aminotransferase were lower than twice the normal value. Creatinine levels were normal in all patients but lactate dehydrogenase was increased in 31 of 67 patients (46%). Twenty-one of 67 patients (31%) showed elevated levels of hypersensitive C-reactive protein. Less common findings were elevated levels of Prothrombin time and creatine kinase, and lymphocytopenia, which was present in all patients. The symptomatic patient group showed a higher proportion of lymphocytopenia than the asymptomatic group. Prealbumin was significantly lower in the group with clinical symptoms compared with asymptomatic patients (241.8±68.2 vs 191.9±60.3, P<0.05)(Table 3).

**Table 3.**
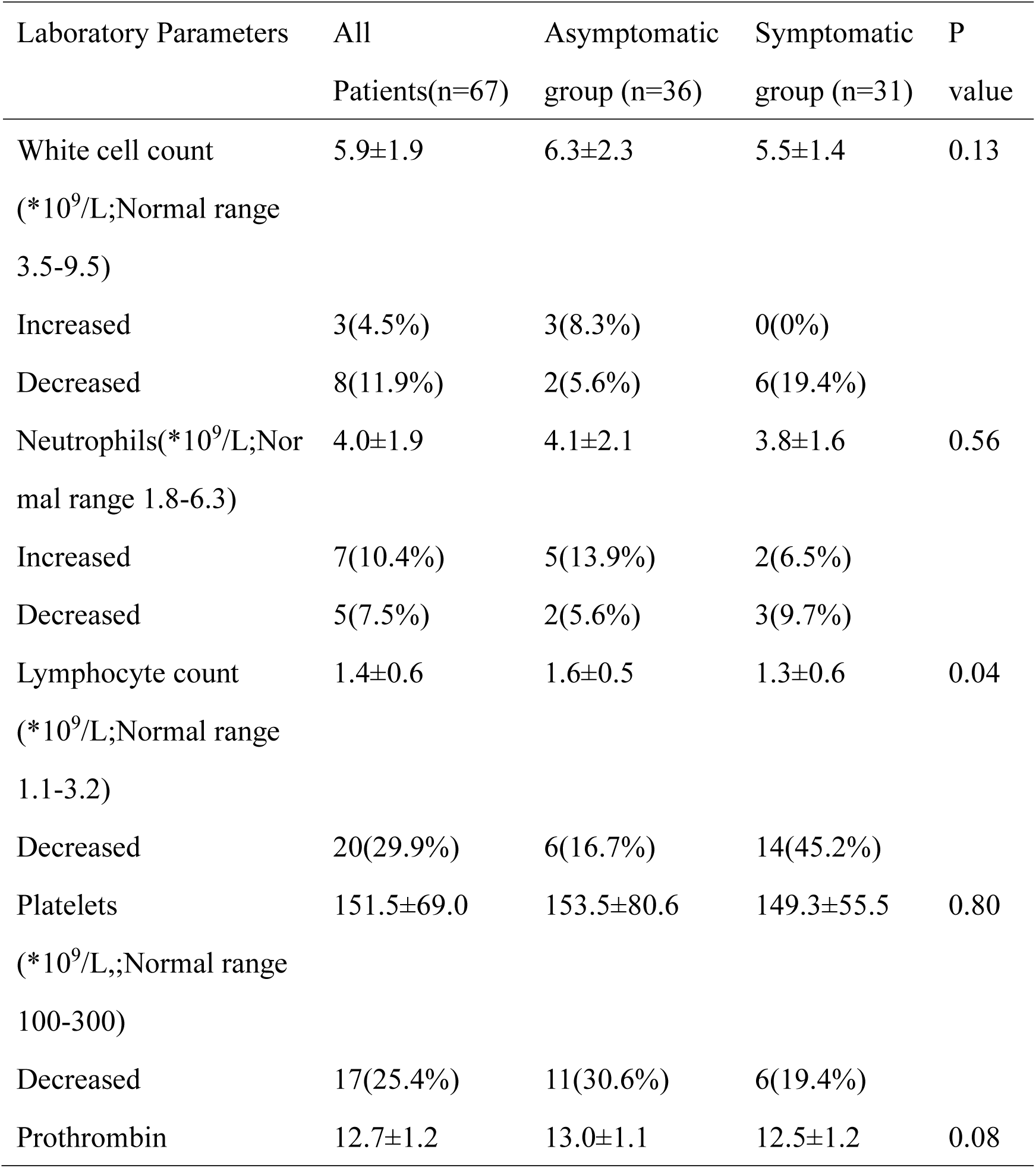
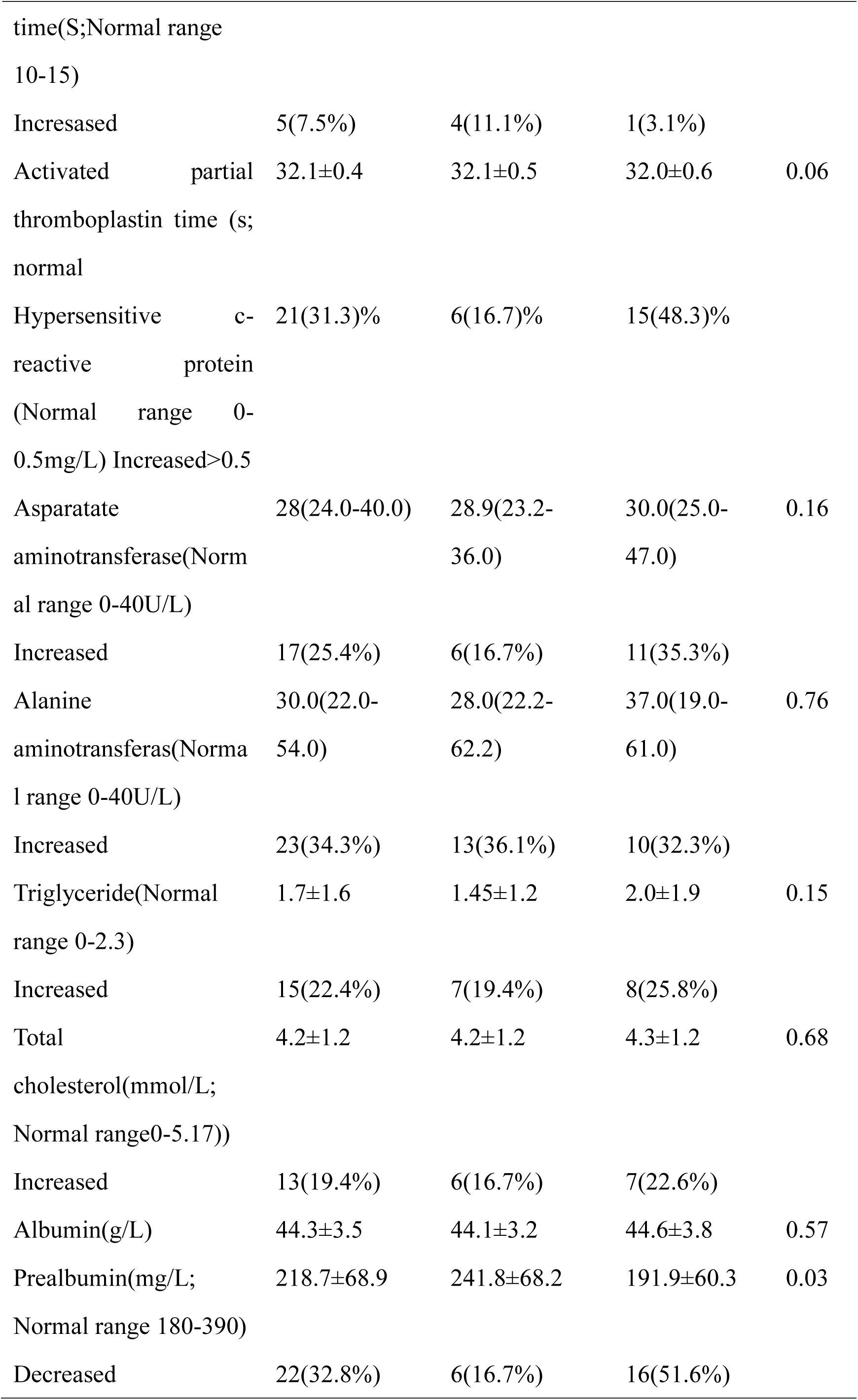
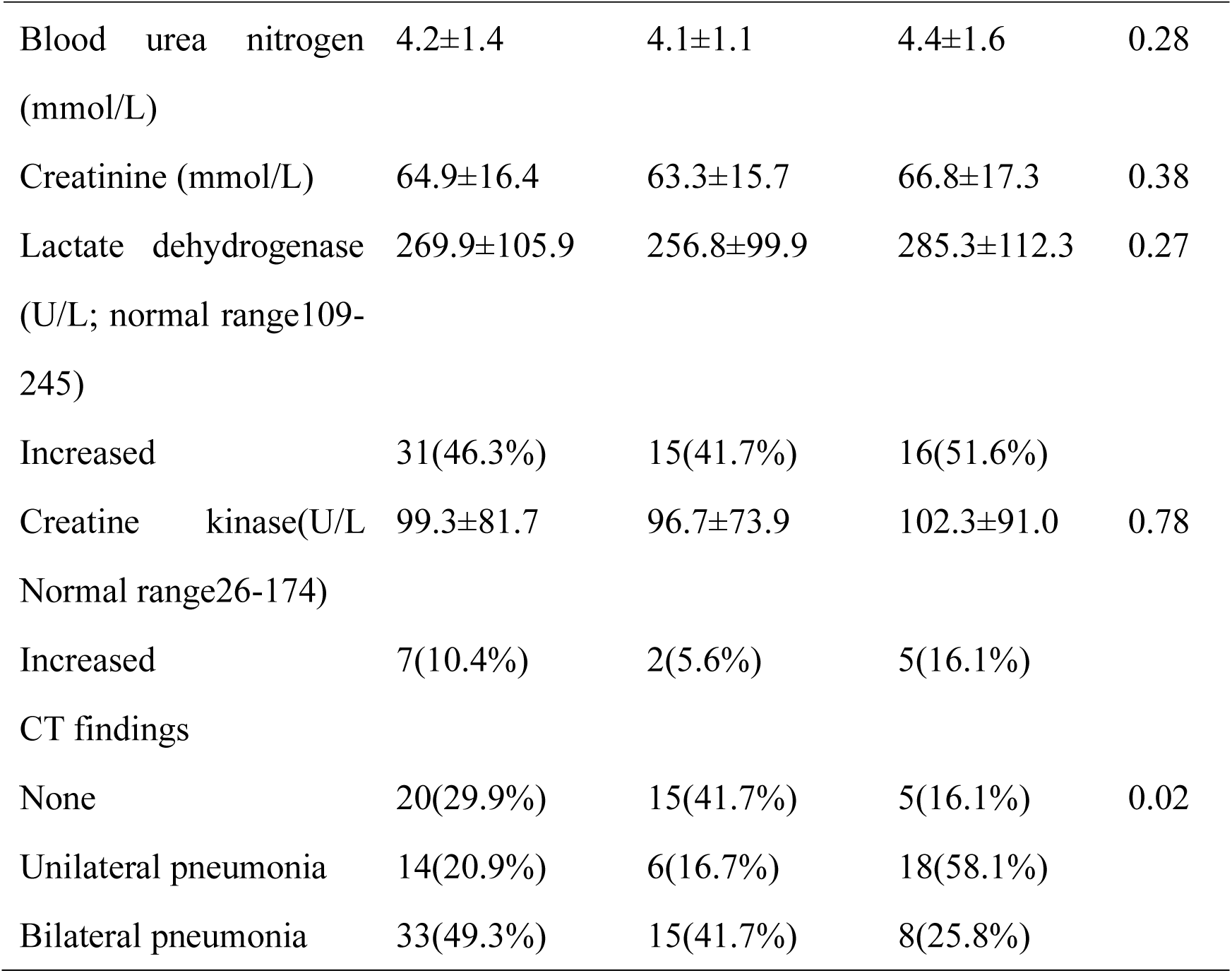
Laboratory results of patients

### Clinical outcomes

By March 5^th^, 52 patients had been discharged, while15 remained in hospital. Severn of 67 had developed to severe disease, requiring treatment at a higher level hospital. The mean age of severe cases were 58.5 years. Three of these severe cases remain in hospital, and their condition has become stable. No patients died of their illness.

## Discussion

Coronavirus disease 2019 (COVID-19) has rapidly spread from Wuhan to other areas of China and beyond. At the beginning of the COVID-19 outbreak, most patients had a history of exposure to COVID-19 cases from Wuhan or had travelled to Wuhan. With advancing time, the medical history associated with case exposure to SARS-CoV-2 infected patients from Wuhan has become less obvious. Human to human transmission is now occurring, resulting in imported cases with no direct contact with patients in Wuhan. In our study, we assessed 67 SARS-CoV-2 infected Tibetan patients living in theQinghai-Tibetan plateau. None of these patients had a history of exposure to COVID-19 cases from Wuhan or had travelled to Wuhan. However, most of them had a history of attending a gathering or contact with a SARS-CoV-2-infected family. Isolation of cases and contact tracing has been implemented for COVID-19^2^. In our study, most patients did not present with typical clinical symptoms at time of admission. This is inconsistent with recent studies, which have most commonly found fever and cough to be the dominant symptoms^3, 4^. In our study, fever was only found in 10% of patients on admission and increased to13.5% during hospitalization. Cough was only presented by 18% of patients. The absence of fever and cough was frequent in our study. Unlike the other studies that included only the cases who actively sought medical attention^3^, our cohort included sub-or preclinical cases identified by local CDC collection of nasal and pharyngeal secretion samples from most residents to detect SARS-CoV-2 nucleic acid, even though the majority of residents had no clinical symptoms or any history of exposure to COVID-19 cases. Through active screening, we found 36 virus carriers without clinical symptoms. If surveillance relies on fever detection or on patients actively seeking medical advice due to clinical symptoms, many potential virus carriers may be missed.

Lymphocytopenia was common, especially in those with clinical symptoms, which is consistent with the data reported recently. Nearly half of patients showed increased levels of LDH, while one-third of patients suffered liver injury and decreased levels of prealbumin. However, these changes were mild. Angiotensin converting enzyme 2 (ACE2) may act as a potential intermediate host receptor which transmitting SARS-CoV-2 to human. ACE2 is expressed in liver tissue and an overactive inflammatory response in patients with SARS-CoV-2 infection may cause increased ACE2 expression, and thus result in the observed liver tissue injury ^5, 6^. Therefore, in addition to the obvious target organ of the lungs, the liver is another important organ that is vulnerable following infection with SARS-CoV-2.

In our study, the condition of most patients was mild, with only 10% of patients developing to severe disease. After antiviral and oxygen therapies, the patients’ conditions gradually stabilized, with none succumbing to disease. The fatality rate was therefore lower than that reported by the national official statistics, which recorded a rate of death of 3100 among 80000 cases of COVID-19 to date in China. One reason for this discrepancy may be that in our study, all the patients were imported cases resulting from serial human to human transmission rather than direct contact with cases in Wuhan. In addition, the altitude at which our cohort lives is higher than 3000 m and so virus viability and virulence may be decreased. The second reason may be that the median age was 39.3 years in our study, which was younger than that reported by Huang et al ^7^. Interestingly, the median age of patients with clinical symptoms was older than that of asymptomatic patients (44.9 vs 34.4 years, P=0.02). In general, older persons appear more susceptible to COVID-19 and more likely to suffer severe disease, which may be due to underlying health issues and comorbidities ^8^. In our study, the mean age of severe cases were 58.5 years. A further reason for the discrepancy in mortality rates between our study and the national figures may be that because the local CDC actively screened a large number of residents, approximately half of the patients were identified and admitted to hospital before clinical symptoms appeared. Patients therefore received treatment at the earliest stages of disease. Early identification and timely treatment are of crucial importance for effective prevention of severe disease.

Our study has some notable limitations. First, only 67 patients were included, although this study describes the largest cohort of Tibetan patients. With the effective measures taken by the government, the number of new patients has decreased.

Second, some patients remained in hospital and the outcome is unknown at the time of data cutoff. We will continue to focus on the prognosis of these patients and report outcomes in due course.

In conclusion, imported cases of SARS-CoV-2 infection in Tibetan patients were generally mild in this high-altitude area. Absence of fever or radiologic abnormalities on initial presentation was common. Our findings highlight the importance of active screening for residents who live in areas with high incidence rate of COVID-19.

## Data Availability

All data was collected and analysed by our team.We would to take responsibility for it.

## Acknowledgements

We thank Gillian Campbell, PhD, from Liwen Bianji, Edanz Group China (www.liwenbianji.cn/ac), for editing the English text of a draft of this manuscript.

